# Pregnancy Cohorts as Sentinel Populations: Evidence From Longitudinal SARS-CoV-2 Serology in Malawi

**DOI:** 10.1101/2025.06.22.25330095

**Authors:** Louise M. Randall, Nicholas Kiernan-Walker, Ernest Moya, Glory Mzembe, Alistair R. D. McLean, Rebecca Harding, Ramin Mazhari, Gomezgani Mhango, Katherine L. Fielding, Ivo Mueller, Martin N. Mwangi, Sabine Braat, Kamija Phiri, Sant-Rayn Pasricha, Emily M. Eriksson, Ricardo Ataíde

## Abstract

1.

**Background:** Despite COVID-19’s global impact, surveillance remains challenging in resource-limited settings. This study investigated IgG to SARS-CoV-2 antigens in Malawian pregnant women participating in the REVAMP trial, where no COVID-19 clinical cases or positive SARS-CoV-2 tests were reported during the pandemic (April 2020 – September 2021).

**Methods:** We analysed serum samples collected from November 2018-September 2021 from 852 pregnant women at enrolment (second trimester) and delivery, measuring IgG against five SARS-CoV-2 antigens using a multiplex bead assay. IgG to Tetanus toxoid served as a positive control, and IgG to four seasonal coronavirus antigens and Influenza A were used for context.

**Findings:** Women recruited after the start of the pandemic were younger than those recruited before the emergence of COVID-19 (median age 19 vs 21 years) and more likely primigravid (60.1% vs 51.1%). IgG levels to SARS-CoV-2 antigens showed sharp increases of 18.5%-29.7% every 30 days during COVID waves 2 and 3. Overall seropositivity to SARS-CoV-2 antigens reached 39.3% during the pandemic; however, the 14.7% seropositivity detected pre-pandemic demonstrates the presence of cross-reactive antibody responses against other antigens in Malawi. Surprisingly, pregnancies within the pandemic showed improved outcomes, with longer gestations (mean difference: 0.6 weeks [95% CI: 0.2-0.9]) and higher birth weights (mean difference: 169.3g, [65.9-272.6]) when comparing to pregnancies occurring pre-pandemic. We found no evidence that SARS-CoV-2 IgG levels were associated with pregnancy outcomes.

**Interpretations:** This study demonstrates that serological surveillance of pregnancy cohorts reveals undetected SARS-CoV-2 exposure where traditional testing was limited. The improvement in pregnancy outcomes during the pandemic suggests community-level exposure did not offset the social impacts of the pandemic in this population. These findings highlight pregnancy cohorts as valuable sentinel populations for infectious disease surveillance, emphasising the importance of serosurveillance in resource-limited settings.

**Funding:** Bill & Melinda Gates Foundation (INV-010612), National Health and Medical Research Council (GNT1158696 and GNT2009047).

**Research in context:** Evidence before this study: The COVID-19 pandemic presented unique surveillance challenges in resource-limited settings where constrained testing capacity complicated efforts to track SARS-CoV-2 transmission. In Malawi, despite early public health measures, significant challenges in disease surveillance left the true extent of viral spread uncertain. We searched Pubmed, Medline and official statistics from the Malawi government for “pregnancy” and “COVID” or “SARS-CoV-2” and “antibody” or “serology” between November 2019 and February 2025. Official data suggested relatively low case numbers across the population compared to other regions; however, limited testing capacity, particularly in rural areas, made it difficult to ascertain actual infection rates. Serological studies across Africa revealed SARS-CoV-2 seroprevalence estimates ranging from 0-63%, with Malawian studies estimating 10-85% depending on the sampling strategy and population.

**Added value of this study:** This study leveraged samples from the REVAMP trial (November 2018-September 2021) to investigate undetected SARS-CoV-2 transmission in a Malawian pregnancy cohort where no clinical cases were reported. The trial provided reliable pre-pandemic baseline samples and consistent monitoring during the pandemic period. Despite the absence of reported symptoms and positive tests as part of the trial, population-level IgG to SARS-CoV-2 antigens increased during well-defined COVID periods. Increased inter-antigenic correlation during the COVID period supports true population transmission. Surprisingly, pandemic pregnancies showed improved outcomes, with longer gestations and higher birth weights, while SARS-CoV-2 IgG levels were not associated with pregnancy outcomes.

**Implications of all the available evidence:** This study provides evidence supporting pregnancy as a sensitive sentinel period for infectious disease surveillance. Our findings demonstrate that serological surveillance can reveal undetected SARS-CoV-2 transmission where traditional testing was limited. The high background seropositivity for SARS-CoV-2 antigens detected in Malawi warrants further research into its clinical significance. Improved pregnancy outcomes during the pandemic, despite serological evidence of transmission, suggest community-level exposure did not adversely affect pregnancy in this population during this period. This may be due to exposure levels below a significant threshold, where SARS-CoV-2’s negative impacts are less significant than the improvements from reductions in other infections, or the infection impacts are outweighed by demographic shifts in the study population. These findings highlight pregnancy cohorts as valuable sentinel populations, emphasising the importance of serosurveillance in resource-limited settings where traditional surveillance methods may be inadequate.

## 2. Introduction

The COVID-19 pandemic presented unique surveillance challenges in resource-limited settings, where constrained testing capacity and limited healthcare access complicated efforts to track SARS-CoV-2 transmission.^1^ In Malawi, despite early public health measures, significant challenges in disease surveillance and response coordination left the true extent of viral spread uncertain. While official statistics suggested low case numbers, compared to other global regions, limited molecular testing capacity and accessibility, particularly in rural areas, meant that actual infection rates remained difficult to ascertain. ^2,3^

This surveillance challenge was strikingly illustrated in the REVAMP trial—a randomized controlled trial of iron supplementation in pregnancy conducted in southern Malawi.^4^ Running from November 2018 through September 2021, the trial spanned pre-pandemic and pandemic periods, including three distinct COVID-19 waves that occurred in Malawi.^5^ Despite regular participant monitoring through at least four study visits between second-trimester enrolment and delivery, no trial participant reported receiving a positive SARS-CoV-2 test or presented with respiratory symptoms indicative of COVID-19. A cross-sectional seroprevalence survey of blood donations at the Malawi Blood Transfusion Service, suggested that the overall seropositivity of SARS-CoV-2 in Malawi in August 2021 could reach 65%.^6^ As such, the absence of reported cases within REVAMP, a closely monitored cohort going through a pandemic, suggested that infections were going undetected.

Serological surveillance offers a powerful tool for addressing such detection challenges, particularly when supported by appropriate validation measures. By determining antibody responses rather than active infections, serosurveillance can reveal historical exposure patterns and provide insights into population-level transmission dynamics that traditional surveillance methods might miss.^7^ The value of this approach has been well-demonstrated in various infectious disease contexts.^8–11^

Pregnancy cohorts are particularly valuable for serosurveillance, as they provide year-round contact points for sample collection and monitoring.^12,13^ For example, in malaria-endemic areas of Africa, serosurveillance of pregnant women is increasingly seen as a reliable and easy way to determine trends in malaria transmission across the population.^10,12,13^ Additionally, pregnancy-specific interventions, such as vaccinations during governmental health programs, create built-in immunological controls that can validate changes seen in incidental serological findings— e.g. tetanus vaccination during pregnancy provides a reliable immunological stimulus that can verify both healthcare delivery and antibody detection methods.^14^

The REVAMP trial presented an ideal platform for investigating potential undetected transmission of SARS-CoV-2 through serosurveillance during pregnancy. Its timeline included reliable pre- and post-pandemic samples from the same population—rare in COVID-19 serological studies—while its regular monitoring schedule ensured consistent sample collection throughout the pandemic period. The trial’s comprehensive data collection on several demographics, clinical outcomes, and healthcare delivery patterns offered multiple avenues for validating the serological findings. We aimed to leverage this unique opportunity to assess evidence of undetected SARS-CoV-2 exposure through validated serological approaches. In addition, we evaluated the impact of changes to SARS-CoV-2 serology on pregnancy outcomes.

## 3. Methods

### Study design

All samples are from the REVAMP trial, which took place in southern Malawi, in the Zomba and Blantyre districts. The trial protocol, statistical analysis plan and primary results have been published.^4,15^ Briefly, pregnant women with ultrasound-confirmed singleton second-trimester pregnancy were individually randomized to receive either an intravenous iron infusion of Ferric Carboxymaltose [Vifor Pharma] or standard of care oral iron if they met the eligibility criteria. Eligibility criteria included haemoglobin of <10.0g/dL (by HemoCue 301+ [Angelholm, Sweden]), and a negative malaria rapid diagnostic test, among others. The primary endpoint of the trial was anaemia prevalence at 36 weeks’ gestation. Venous blood samples were taken throughout the pregnancy, including at enrolment (13-26 weeks’ gestation), and delivery. Blood was processed within two hours of collection and plasma was aliquoted and stored locally at −80 °C until being shipped to Melbourne, Australia. All women were eligible to receive pregnancy immunisations from the Malawi Government, following the national Expanded Programme of Immunisation strategy. These include a dose of Tetanus toxoid (Tt) at the first antenatal visit (often the time of first contact with the REVAMP trial team), a second dose 4 weeks after the first dose and a third dose 6 months after the second dose.^16^

The trial received ethics approvals from the College of Medicine, University of Malawi, Malawi (P.02/18/2357), and The Walter and Eliza Hall Institute (WEHI), Australia (18/02) and was prospectively registered (ACTRN12618001268235). An independent data and safety monitoring board oversaw the trial. Secondary analyses of the trial samples were included in the study protocol.

### COVID-19 Setting

All dates related to the COVID-19 pandemic were obtained from publicly available Malawian Government sources.^5^ The first official case of COVID-19 in Malawi was reported on April 2^nd^ 2020, and marked the beginning of the first wave of infections (Figure 1).^5^ Following the establishment of containment policies, and at the request of the Malawi Ministry of Health, REVAMP amended its protocol and adopted COVID measures in May of 2020 (i.e. limiting the number of contact visits between the team and the participants, as well as limiting the duration of those visits).^15^ In January 2021, the Malawi government implemented a national lockdown. Vaccinations were introduced in Malawi on March 11^th^ 2021, with the ChAdOx1-S COVID-19 vaccine being delivered.^5^ Study participants were deliberately not asked whether they had received COVID-19 vaccines as there was a lot of misconception, and it was thought that this question could affect retention in subsequent visits. However, at each visit, participants were asked to report any concomitant medication they were on or any prior visits to a health centre, and we had no indication that any of the study participants received the vaccination. Malawi experienced three waves of COVID-19 during the lifespan of REVAMP.^5^ During the entire trial, there were no participant self-reported nor trial team-confirmed COVID-19 cases within the cohort, and there were no observed or reported positive SARS-CoV-2 tests.

**Figure 1:**
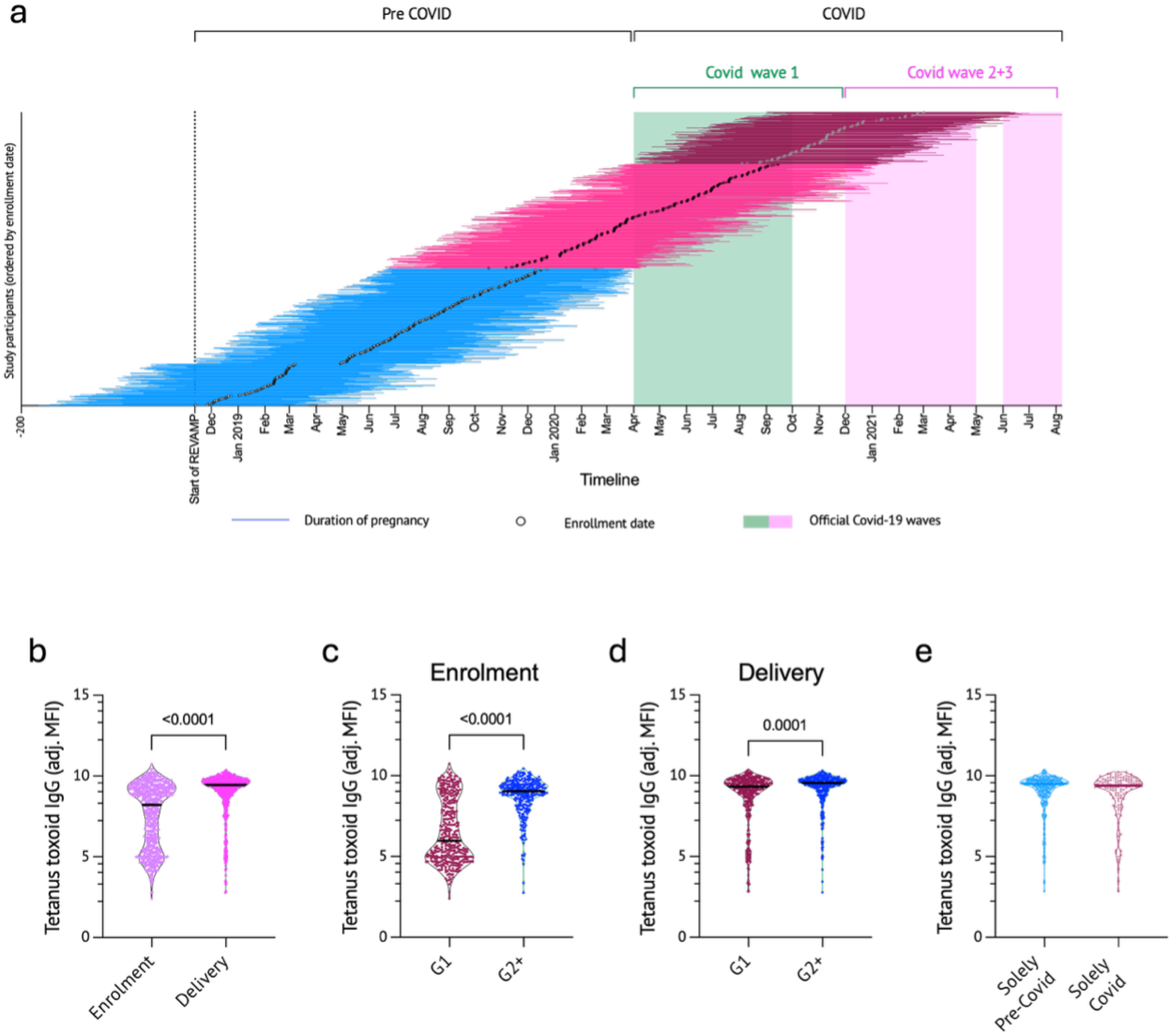
Participant timelines and cohort characteristics across pre-pandemic and pandemic periods. (a) Individual participant timeline distribution across the study period. Horizontal lines represent each participant’s pregnancy duration, with enrollment dates marked as circles. Blue horizontal lines are pregnancies that occurred exclusively in the pre-COVID-19 period. Magenta lines are pregnancies that occurred partially in the COVID-19 period. Dark red lines are pregnancies entirely occurring within the COVID-19 period. Shaded boxes represent reported COVID-19 waves (wave 1 – green; waves 2 and wave 3 – pink). (b) Anti-tetanus toxoid IgG levels by visit date (enrolment and delivery) (unpaired Welch’s t-test). (c) Anti-tetanus toxoid IgG levels at enrollment between first-time pregnancies (G1) and second or higher pregnancies (G2+) (unpaired Welch’s t-test). (d) anti-tetanus toxoid IgG levels at delivery between first-time pregnancies (G1) and second or higher pregnancies (G2+) (unpaired t-test). (e) Anti-tetanus toxoid IgG levels between pregnancies occurring entirely within the pre-COVID period (solely pre-COVID) or pregnancies that occurred exclusively during the Malawi COVID-19 period (Solely COVID) (unpaired t-test). All serological values are shown as log_e_-transformed median fluorescence intensity (MFI).

### Serological multiplex bead-array

A multiplex serological assay was used to detect antigen-specific IgG antibodies targeting the SARS-CoV-2 Spike protein, Spike-derived antigens (S1, S2 and receptor binding domain (RBD)) and nucleoprotein (NP), as previously described.^17^ Additionally, we assessed antibodies to seasonal coronaviruses antigens (NL63, OC43, HKU1 and 229E) and Influenza virus A (H1N1). Antibodies to Tt were also measured as an immunological positive control.^16^ Briefly, individual plasma samples were incubated with fluorescent magnetic beads coated with the relevant antigens. Antigen-specific antibodies were then identified using a secondary anti-human IgG Fc antibody conjugated to phycoerythrin (PE) and analyzed with the MAGPIX® system. All samples were run with the analysts blinded to the participants’ characteristics.

### Statistical analysis

The sample size was determined by the available data of all randomised women with a plasma sample at study enrolment or delivery. Baseline characteristics were summarised and compared before and after COVID-19 emergence in Malawi using either two-sample t or Wilcoxon rank-sum test for continuous data or chi-square or Fisher’s exact test for categorical data. Antibody levels were quantified as adjusted mean fluorescent intensity (aMFI) derived from standard curves. Seropositivity for SARS-CoV-2 antibodies was determined as IgG levels exceeding the mean plus two standard deviations from pre-pandemic pregnancies. Longitudinal log_e_-transformed IgG levels were visualised using locally weighted (LOWESS) regression with restricted cubic spline curve, and 10 smoothing points.^18^ Linear regression models with two inflexion points (start of the COVID-19 pandemic in Malawi - April 2nd 2020; beginning of the second COVID-19 wave - January 1st 2021), resulted in three time segments. Models accounted for variables assumed to be prognostic or predictive of the outcomes: maternal age, body mass index, primiparity, sex of the newborn, HIV status, income source and treatment group. Estimates and confidence intervals (CI) were back-transformed to reflect a per cent of change over 30 days.

Linear regression models were used to quantify associations between pregnancy outcomes of interest (gestational age, birth weight and low birth weight) and serological data at delivery as exposures with adjustment for confounding and prognostic variables - maternal age, primigravidity, body mass index, HIV status, and treatment group. Spearman correlations assessed the IgG relationships during the Pre-COVID or COVID periods. No adjustment for multiple testing was done.

Small for gestational age was defined as a birthweight below the 10^th^ percentile for gestational age according to INTERGROWTH-21 standards;^19^ Low birth weight is defined as a birth weight <2500 g; Premature birth is a birth occurring at <37 weeks’ gestation; Foetal loss represents a composite of pregnancy loss and stillbirth. All statistical analyses were conducted in Stata v18.0 (StataCorp, College Station, TX) and GraphPad Prism v10 (GraphPad Software, San Diego, CA, USA).

## 4. Results

### Trial Population reflected COVID-19 impacts

The REVAMP trial recruited 862 participants between November 2018 and March 2021, with the last delivery in the trial occurring in September 2021, thus pregnancies in REVAMP spanned pre-pandemic and pandemic periods (Figure 1a). A total of 852/862 (98.8%) women had available serum samples and were included in this nested study (Table 1), including 147 samples from enrolment only, 30 samples from delivery only, and 675 samples for both enrolment and delivery. The baseline characteristics of the women with a sample analysed at only one time point were similar to women who had samples analysed at both time points (Table S1).

**Table 1-.**
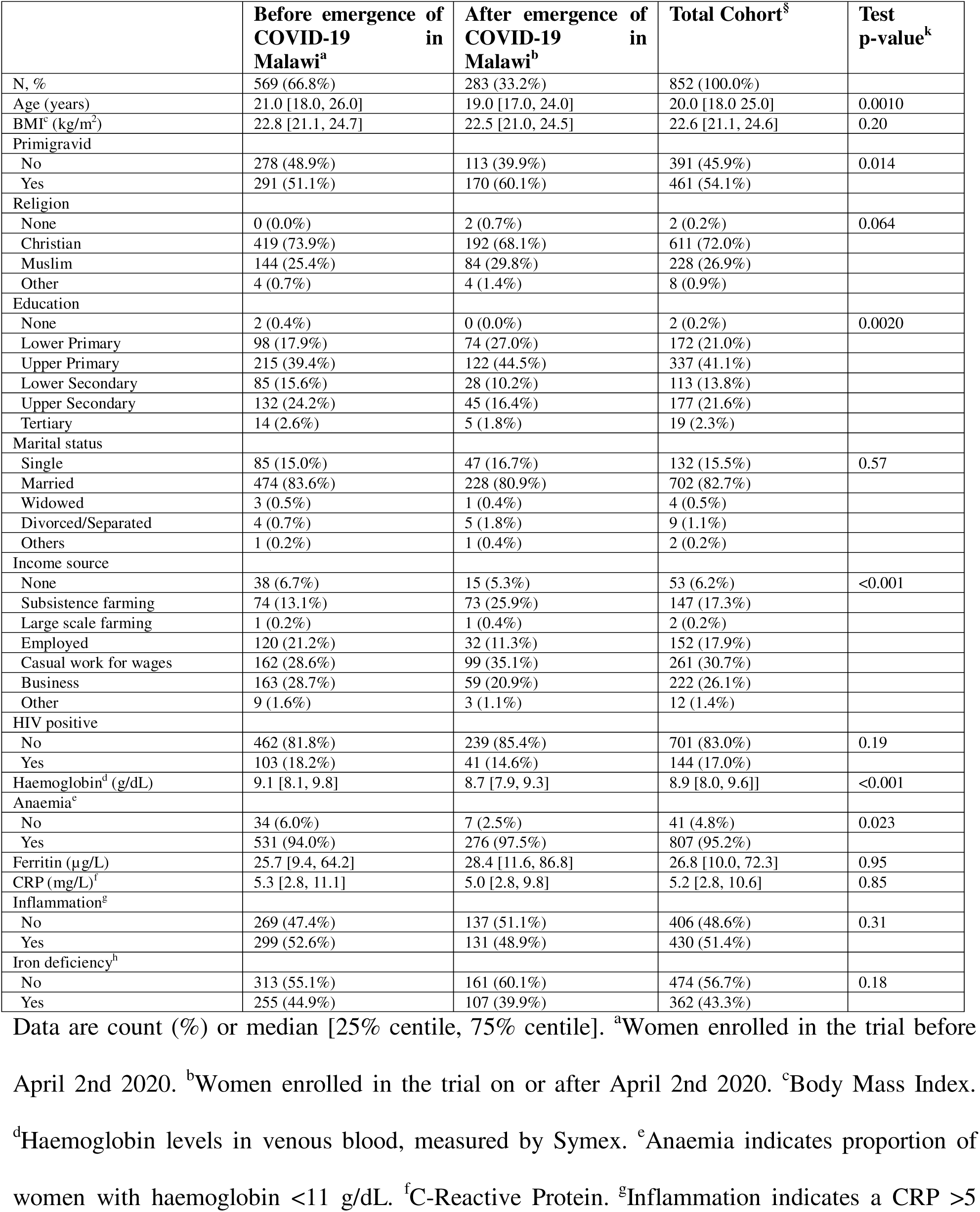
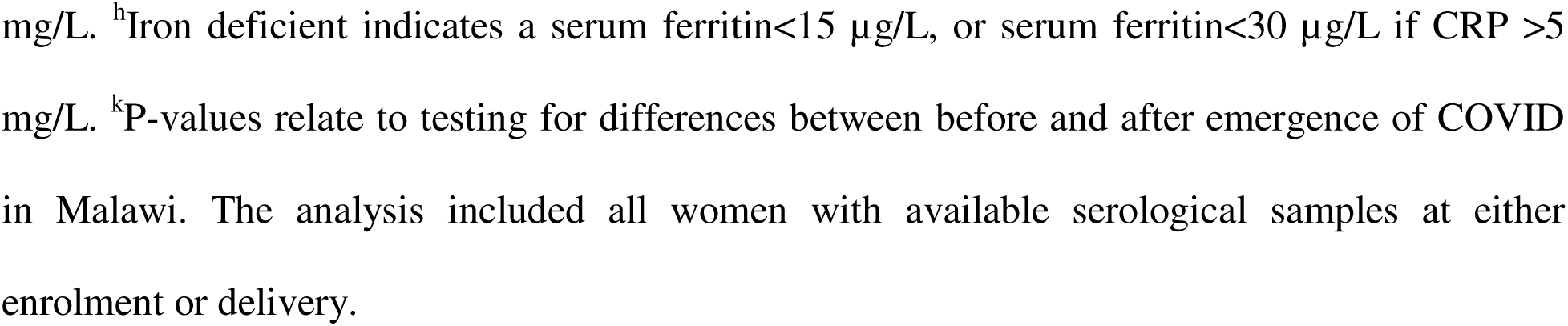
Baseline characteristics according to the timing of the emergence of COVID-19 in Malawi.

|  | Before emergence of COVID-19 in Malawi<sup>a</sup> | After emergence of COVID-19 in Malawi<sup>b</sup> | Total Cohort<sup>§</sup> | Test p-value<sup>k</sup> |
| --- | --- | --- | --- | --- |
| N, % | 569 (66.8%) | 283 (33.2%) | 852 (100.0%) |  |
| Age (years) | 21.0 [18.0, 26.0] | 19.0 [17.0, 24.0] | 20.0 [18.0, 25.0] | 0.0010 |
| BMI<sup>c</sup> (kg/m<sup>2</sup>) | 22.8 [21.1, 24.7] | 22.5 [21.0, 24.5] | 22.6 [21.1, 24.6] | 0.20 |
| Primigravid |  |  |  |  |
| No | 278 (48.9%) | 113 (39.9%) | 391 (45.9%) | 0.014 |
| Yes | 291 (51.1%) | 170 (60.1%) | 461 (54.1%) |  |
| Religion |  |  |  |  |
| None | 0 (0.0%) | 2 (0.7%) | 2 (0.2%) | 0.064 |
| Christian | 419 (73.9%) | 192 (68.1%) | 611 (72.0%) |  |
| Muslim | 144 (25.4%) | 84 (29.8%) | 228 (26.9%) |  |
| Other | 4 (0.7%) | 4 (1.4%) | 8 (0.9%) |  |
| Education |  |  |  |  |
| None | 2 (0.4%) | 0 (0.0%) | 2 (0.2%) | 0.0020 |
| Lower Primary | 98 (17.9%) | 74 (27.0%) | 172 (21.0%) |  |
| Upper Primary | 215 (39.4%) | 122 (44.5%) | 337 (41.1%) |  |
| Lower Secondary | 85 (15.6%) | 28 (10.2%) | 113 (13.8%) |  |
| Upper Secondary | 132 (24.2%) | 45 (16.4%) | 177 (21.6%) |  |
| Tertiary | 14 (2.6%) | 5 (1.8%) | 19 (2.3%) |  |
| Marital status |  |  |  |  |
| Single | 85 (15.0%) | 47 (16.7%) | 132 (15.5%) | 0.57 |
| Married | 474 (83.6%) | 228 (80.9%) | 702 (82.7%) |  |
| Widowed | 3 (0.5%) | 1 (0.4%) | 4 (0.5%) |  |
| Divorced/Separated | 4 (0.7%) | 5 (1.8%) | 9 (1.1%) |  |
| Others | 1 (0.2%) | 1 (0.4%) | 2 (0.2%) |  |
| Income source |  |  |  |  |
| None | 38 (6.7%) | 15 (5.3%) | 53 (6.2%) | <0.001 |
| Subsistence farming | 74 (13.1%) | 73 (25.9%) | 147 (17.3%) |  |
| Large scale farming | 1 (0.2%) | 1 (0.4%) | 2 (0.2%) |  |
| Employed | 120 (21.2%) | 32 (11.3%) | 152 (17.9%) |  |
| Casual work for wages | 162 (28.6%) | 99 (35.1%) | 261 (30.7%) |  |
| Business | 163 (28.7%) | 59 (20.9%) | 222 (26.1%) |  |
| Other | 9 (1.6%) | 3 (1.1%) | 12 (1.4%) |  |
| HIV positive |  |  |  |  |
| No | 462 (81.8%) | 239 (85.4%) | 701 (83.0%) | 0.19 |
| Yes | 103 (18.2%) | 41 (14.6%) | 144 (17.0%) |  |
| Haemoglobin<sup>d</sup> (g/dL) | 9.1 [8.1, 9.8] | 8.7 [7.9, 9.3] | 8.9 [8.0, 9.6] | <0.001 |
| Anaemia<sup>e</sup> |  |  |  |  |
| No | 34 (6.0%) | 7 (2.5%) | 41 (4.8%) | 0.023 |
| Yes | 531 (94.0%) | 276 (97.5%) | 807 (95.2%) |  |
| Ferritin (µg/L) | 25.7 [9.4, 64.2] | 28.4 [11.6, 86.8] | 26.8 [10.0, 72.3] | 0.95 |
| CRP (mg/L)<sup>f</sup> | 5.3 [2.8, 11.1] | 5.0 [2.8, 9.8] | 5.2 [2.8, 10.6] | 0.85 |
| Inflammation<sup>g</sup> |  |  |  |  |
| No | 269 (47.4%) | 137 (51.1%) | 406 (48.6%) | 0.31 |
| Yes | 299 (52.6%) | 131 (48.9%) | 430 (51.4%) |  |
| Iron deficiency<sup>h</sup> |  |  |  |  |
| No | 313 (55.1%) | 161 (60.1%) | 474 (56.7%) | 0.18 |
| Yes | 255 (44.9%) | 107 (39.9%) | 362 (43.3%) |  |

Data are count (%) or median [25% centile, 75% centile]. <sup>a</sup>Women enrolled in the trial before April 2nd 2020. <sup>b</sup>Women enrolled in the trial on or after April 2nd 2020. <sup>c</sup>Body Mass Index. <sup>d</sup>Haemoglobin levels in venous blood, measured by Symex. <sup>e</sup>Anaemia indicates proportion of women with haemoglobin <11 g/dL. <sup>f</sup>C-Reactive Protein. <sup>g</sup>Inflammation indicates a CRP >5
mg/L. <sup>h</sup>Iron deficient indicates a serum ferritin<15 µg/L, or serum ferritin<30 µg/L if CRP >5 mg/L. <sup>k</sup>P-values relate to testing for differences between before and after emergence of COVID in Malawi. The analysis included all women with available serological samples at either enrolment or delivery.

A total of 569 (66.8%) women were enrolled before and 283 (33.2%) women were enrolled after April 2^nd^ 2020, the date of the first case of COVID-19 registered in Malawi (Table 1) while 358 (42.0%) pregnancies were spent solely under the Pre-COVID period and 135 (15.8%) solely under the COVID periods (Figure 1a).

We observed differences in demographic characteristics of the trial participants before and after the emergence of COVID-19 in Malawi (Table 1). The impact of time of recruitment on demographics revealed maternal age decreased across the period of the trial, from a median of 21 years (IQR [19-27]) in the pre-COVID period to 19 years (IQR [17-23]) (P=0.0010) during the COVID period. Consistent with this, we observed an increase in the percentage of women in their first pregnancy towards the end of the trial (51.1% (291/569) pre-COVID vs 60.1% (170/283) COVID; P=0.014). Income source also changed significantly from the pre-COVID to the COVID periods (P<0.001), with more women reporting practicing Subsistence farming (13.1% (74/567) pre-COVID vs 25.9% (73/282) COVID) and less women reporting being Employed (21.2% pre-COVID vs 11.4% COVID).

### Serological outcomes validation – Tetanus toxoid

We measured antibodies to Tetanus toxoid (Tt) as a validation tool for our serological assays. As was expected from Malawi’s vaccination policy,^16^ IgG toward Tt were higher at delivery versus enrolment (Figure 1b), and in women with two or more pregnancies compared to women in their first pregnancy – both at enrolment and, though to a lesser degree, at delivery (Figure 1c-d). The overall levels of anti-Tt antibodies measured at delivery were unaffected in those pregnancies carried solely within the COVID period vs those solely in the pre-COVID period (Figure 1e).

### Serological outcomes – SARS-CoV-2

Antibodies to all five of SARS-CoV-2 antigens increased from the beginning of 2021, coinciding with the emergence of COVID waves 2 and 3 in Malawi (Figure 2a and Supplementary figure 1). A wide range of IgG levels to SARS-CoV-2 antigens were present even before the emergence of the COVID-19 pandemic, indicating the occurrence of cross-reactive immune responses in this population. In general, there was a subtle positive trend in antibody levels over time, with values remaining relatively stable through to the end of COVID wave 1, followed by an increase through COVID waves 2 and 3. Linear modelling of this relationship revealed a significant increase in antibodies to SARS-CoV-2 antigens during COVID waves 2 and 3 (Figure 2b, Supplementary Table 2). This increase was also noticeable when comparing mean antibody levels in those pregnancies carried solely within the COVID period versus those occurring in the pre-COVID (Figure 2c). In addition, the correlations between IgG levels to all SARS-CoV-2 antigens also significantly increased from those pregnancies entirely in the pre-COVID period to those entirely in the COVID period (Figure 2e-d).

**Figure 2:**
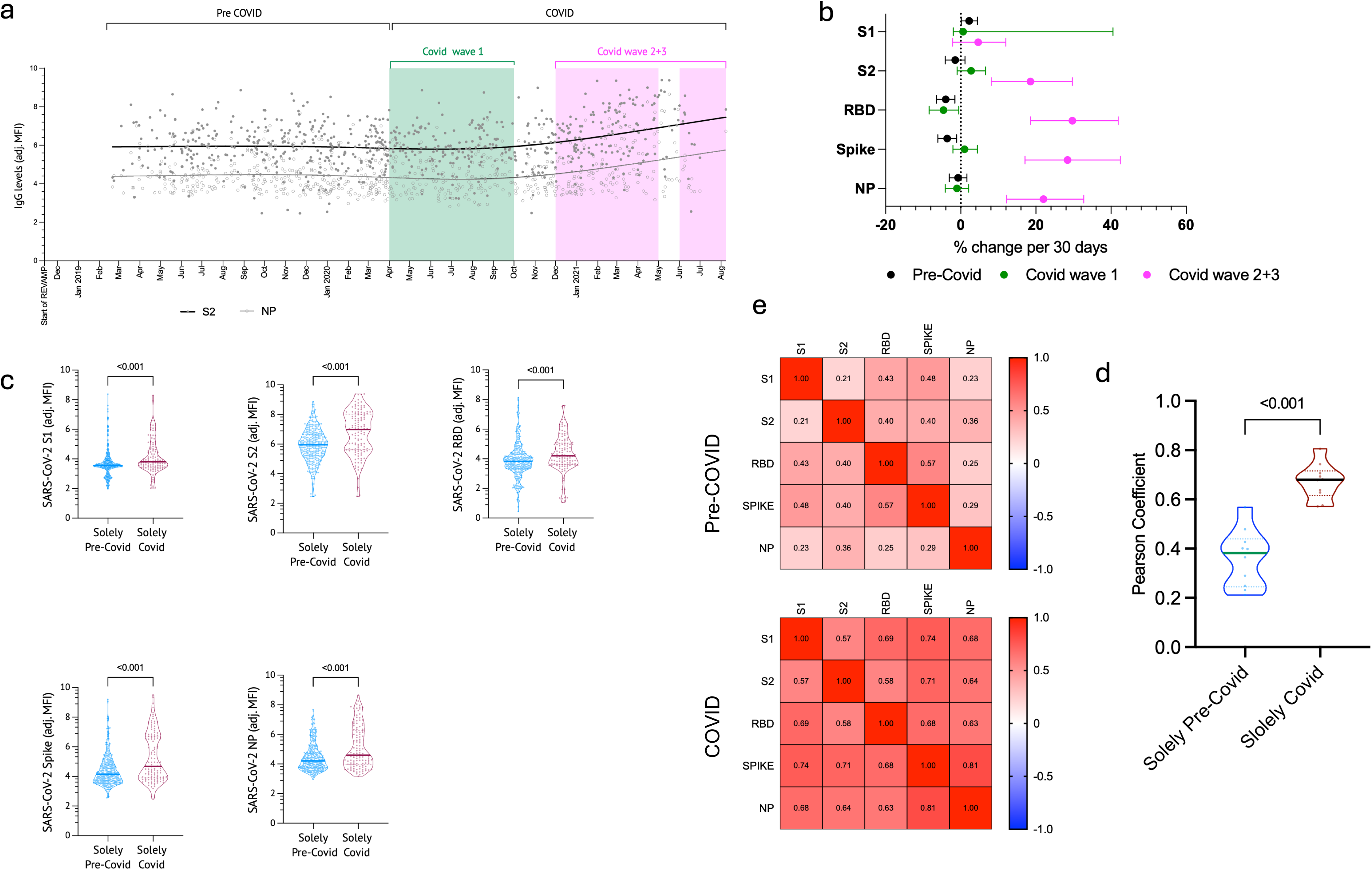
Serological patterns of anti-SARS-CoV-2 antibodies during pregnancy. (a) LOWESS smoothing curves illustrating the relationship between time and anti-SARS-CoV-2 antibodies is shown for SARS-CoV-2 S2 and SARS-CoV-2 NP antigens. The analysis was conducted using GraphPad Prism v10. This method fits a non-parametric regression and makes no assumptions about the functional form of the relationship. The graph includes the original data points as scatter markers to allow assessment of the density and distribution of observations across the range of values. (b) Mean percent change in IgG levels per 3 days across the three time periods assessed after fitting a linear regression model. (c) Comparison of mean antibody levels to SARS-CoV-2 antigens between pregnancies occurring outside (entirely pre-COVID) or inside (entirely COVID) the COVID period (unpaired T-test). (d) Correlation coefficient matrix between antibodies measured against SARS-CoV-2 antigens in samples from pregnancies outside (Solely pre-COVID - top) or inside (Solely COVID - bottom) the COVID period. (e) Comparison of mean correlation coefficient between antibodies against SARS-CoV-2 antigens between pregnancies occurring outside (Solely pre-COVID) or inside (Solely COVID) the COVID period (unpaired T-test).

A total of 102 samples (17.3%) were seropositive for at least one of the SARS-CoV-2 antigens (ranging from 4.3% for RBD to 9.1% against SPIKE). Half of the 102 seropositive samples (49%) were seropositive for 1 of the antigens and only 5 samples (4.1%) were seropositive for all five SARS-CoV-2 antigens. The overall seroprevalence calculated for those pregnancies entirely outside of the COVID-19 period in Malawi was 14.7% (48/327), revealing a significant number of cross-reactive responses to other antigens. In pregnancies occurring entirely within the COVID period, the overall seroprevalence was 39.3% (48/122).

### Serological outcomes – Seasonal coronaviruses and Influenza A

Antibodies to seasonal viruses presented a distinct pattern across the timeline of the trial (Figure 3a-b and Supplementary figure 1). LOWESS visualization showed little variation across the entirety of the trial (Figure 3a). Linear modelling over the three-segments of the trial revealed a general stability over the pre-COVID period— maximum variation was for IgG towards Coronavirus HKU1 (mean % change over 30 days: −4.1% [-5.8, −2.3]) (Supplementary Table 2). This was followed by a small increase in IgG levels during the second period, and an overall decrease during the last period (Figure 3b, Supplementary Table 2). However, the levels of antibodies to these common human coronaviruses, including types NL63, OC43, and HKU1, and to Influenza A (H1N1) remained relatively unchanged when analysing those pregnancies occurring outside or inside the COVID period, except for antibodies to Coronavirus 229E that declined significantly at delivery between solely pre-COVID and solely COVID pregnancies (Figure 3c). There was no significant change to the correlations between antigens to the common coronaviruses and Influenza A between COVID periods (Figure 3c-d).

**Figure 3:**
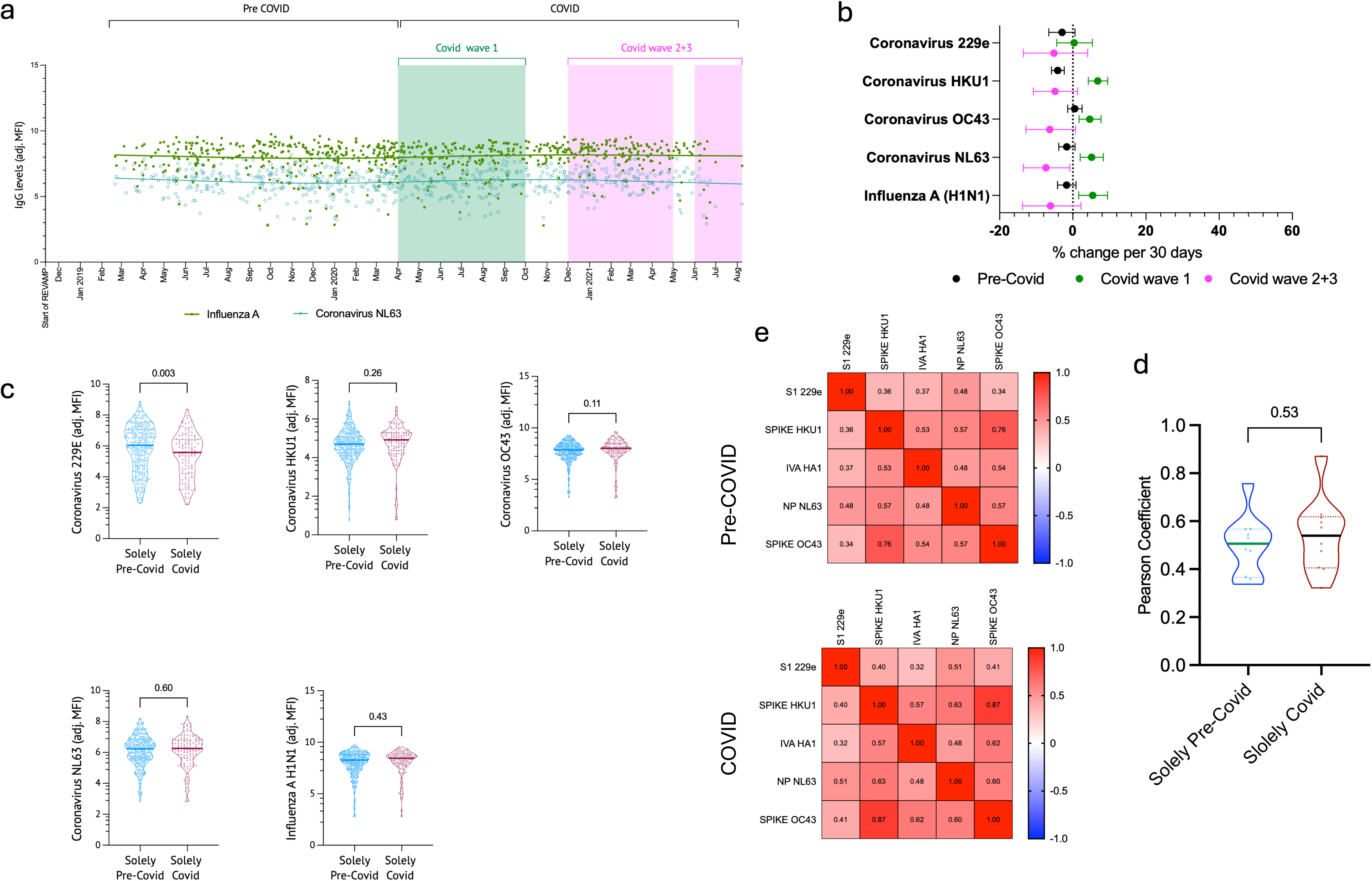
Serological patterns of anti-common Coronavirus and Influenza A (H1N1) antibodies during pregnancy. (a) LOWESS smoothing curves illustrating the relationship between time and antibodies against Influenza A (green) and Coronavirus NL63 (teal) antigens. The analysis was conducted using GraphPad Prism v10. This method fits a non-parametric regression and makes no assumptions about the functional form of the relationship. The graph includes the original data points as scatter markers to allow assessment of the density and distribution of observations across the range of values. (b) Mean percent change in IgG levels per 30 days across the three time periods assessed after fitting a linear regression model. (c) Comparison of mean antibody levels to SARS-CoV-2 antigens between pregnancies occurring outside (entirely pre-COVID) or inside (entirely COVID) the COVID period (unpaired T-test). (d) Correlation coefficient matrix between antibodies measured against SARS-CoV-2 antigens in samples from pregnancies outside (Solely pre-COVID - top) or inside (Solely COVID - bottom) the COVID period. (e) Comparison of mean correlation coefficient between antibodies against SARS-CoV-2 antigens between pregnancies occurring outside (Solely pre-COVID) or inside (Solely COVID) the COVID period (unpaired T-test)

### Pregnancy outcomes of the trial according to COVID period and association with SARS-CoV-2 antibodies

Throughout the trial, and after accounting for confounding variables, gestation duration increased significantly between pregnancies that delivered during the COVID period compared to pre-COVID (adjusted mean difference 0.5 weeks [0.2-0.8]) (Table 2). This difference was still observed when looking at women with pregnancies entirely in the COVID period versus entirely in the pre-COVID period (mean difference 0.6 weeks [0.2, 0.9]). Accordingly, babies had higher birth weights when delivered during the COVID period compared with those delivered during the pre-COVID period (mean difference 69.4 g [-2.4-141.1]), although this increased birthweight was only significant when comparing those pregnancies occurring solely within the COVID period versus occurring solely in pre-COVID (mean difference 169.3 g [65.9, 272.6]). Low birth weight decreased during the COVID period (Odds Ratio (OR) COVID vs pre-COVID: 0.57 [0.37-0.89]), and in those pregnancies solely occurring during COVID versus in pre-COVID (OR Solely COVID vs pre-COVID: 0.40 (0.20-0.77)) (Table 2).

**Table 2.** Effects of COVID-19 period on Pregnancy and Birth Outcomes.

| **Delivery inside vs outside COVID** | **Pre-Covid** | **Covid** | **OR [95% CI] or mean difference [95% CI]** | **P-value** |
| --- | --- | --- | --- | --- |
| Small for gestational age n/N (%) | 146/477 (30.6%) | 92/266 (34.6%) | 1.10 [0.79-1.53] | 0.57 |
| Low birth weight (<2,500g) n/N (%) | 89/484 (18.4%) | 32/265 (12.1%) | 0.57 [0.37-0.89] | 0.013 |
| Premature birth (<37 weeks) n/N (%) | 43/489 (8.8%) | 17/270 (6.3%) | 0.70 [0.39-1.26] | 0.23 |
| Fetal loss n/N (%) | 11/522 (2.1%) | 5/275 (1.8%) | 0.95 [0.32-2.82] | 0.93 |
| Gestation duration (weeks) Mean (SD)<sup>#</sup> | 39.3 (2.1) | 39.8 (1.9) | 0.49 [0.20-0.78] | 0.0010 |
| Birth weight (grams) Mean (SD)<sup>\$</sup> | 2889.9 (501.4) | 2930 (477.8) | 69.36 [-2.39-141.12] | 0.06 |
| **Entire gestation period inside vs outside COVID** |  |  |  |  |
| Small for gestational age n/N (%) | 94/335 (28.1%) | 40/130 (30.8 %) | 0.97 [0.62-1.53] | 0.90 |
| Low birth weight (<2,500g) n/N (%) | 64/340 (18.8%) | 12/127 (9.4%) | 0.40 [0.20-0.77] | 0.0070 |
| Premature birth (<37 weeks) n/N (%) | 35/343 (10.2%) | 8/130 (6.2%) | 0.59 [0.26-1.33] | 0.20 |
| Fetal loss n/N (%) | 5/350 (1.4%) | 1/132 (0.8%) | 0.53 [0.05-5.19] | 0.59 |
| Gestation duration (weeks) Mean (SD)<sup>†</sup> | 39.2 (2.2) | 39.8 (1.6) | 0.6 [0.2-0.9] | 0.0030 |
| Birth weight (grams) Mean (SD)<sup>‡</sup> | 2883.2 (516.3) | 3001.5 (487.4) | 169.3 [65.9-272.6] | 0.0010 |

**Note:** Small for gestational age defined as a birthweight below the 10th percentile for gestational
age according to INTERGROWTH-21 standards;<sup>19</sup> Low birth weight is defined as a a birth
weight < 2500 g; Premature birth is a birth occurring at <37 weeks gestation; Fetal loss
represents a composite of pregnancy loss and stillbirth. <sup>#</sup> N=489 Pre-COVID and 270 COVID.
<sup>\$</sup>N= 484 Pre-COVID and 265 COVID <sup>†</sup>N=343 Pre-COVID and 130 COVID. <sup>‡</sup>N= 340 Pre-
COVID and 127 COVID. OR - Odds Ratio; CI - Confidence Interval. All models controlled for
treatment group, maternal age, body mass index, primigravid status, and HIV status. Analysis
were run on all samples with non-missing data.

In women whose pregnancies occurred within the COVID period, we found no evidence of a statistically significant association between seropositivity to SARS-CoV-2 antigens and pregnancy outcomes. However, the estimates around the associations had very large 95% CI, which is compatible with large effects in either direction (Supplementary Table 3).

## 5. Discussion

In this study we assessed the sensitivity of serosurveillance conducted in pregnancy cohorts to detect longitudinal patterns of pathogen exposure. Using SARS-CoV-2 as an example pathogen, we used samples from the REVAMP clinical trial to examine serological patterns and pregnancy outcomes before and during the COVID-19 pandemic in a well-characterized cohort of Malawian pregnant women. We found that, despite a lack of clinical symptoms and laboratory-confirmed positive tests, levels of antibodies to key antigens of SARS-CoV-2 significantly increased in the population, as did the overall seropositivity prevalence, from a background estimate of 14.7% seropositivity before COVID to an overall seropositivity of 39.3% for pregnancies within the COVID period. Over the full span of the trial, we observed an increase in the mean gestation duration and birth weight of the babies, accompanied by a decrease in the overall prevalence of low birth weight. However, we found no evidence that exposure to SARS-CoV2—as measured by IgG to 5 different antigens — was associated with pregnancy and birth outcomes.

The REVAMP trial commenced in November 2018 and had its last delivery in September 2021. This resulted in continuous sampling throughout pregnancy and up to delivery for women before the emergence of the COVID-19 pandemic and up to the height of the 2021 COVID-19 waves in Malawi. We observed significant demographic shifts throughout the trial’s duration. Women enrolled during the pandemic were significantly younger than those enrolled in the pre-COVID period. This change was accompanied by significant changes to the parity, education and income source measures of the trial cohort—reflecting the younger population. This shift in population age observed during our trial timeline may reflect the broader socio-economic changes that occurred during the pandemic. Malawi reported a decrease in total health service attendance of up to 10% from April 2020 to December 2021, with antenatal services specifically falling by 3%.^20^ It is reasonable to assume that, as lockdowns, school closures and other containment measures were enforced by the Malawian government,^5^ women with young children may have had responsibilities that prevented them from either participating in the study or engage with the health services. These findings also highlight the importance of understanding the characteristics of the underlying population when the time periods of study are lengthy.

Our serological findings contribute to understanding immune responses to SARS-CoV-2 in African pregnant women. A meta-analysis of the seroprevalence of anti-SARS-CoV-2 antibodies across Africa (excluding Malawi) revealed seroprevalences ranging from 0.0-63.0%,^21^ while studies in Malawi, estimated prevalences at between 10-85% depending on the sampling strategy and the population.^6,22,23^ In REVAMP, we measured a peak seroprevalence of 39.3%. Although seropositivity to SARS-CoV-2 does not immediately follow from increases in seroprevalence to single antigens, the increase of antibody levels to all five SARS-CoV-2 antigens during the COVID-19 period, coupled with stronger inter-antigenic correlations, suggests true community transmission occurred in the trial population during this period. This timing agrees with SARS-CoV-2 seroprevalence data obtained from blood donations surveyed from 4 distinct areas of Malawi^6^ and from pregnant women at first antenatal clinic, in a country-wide study which started just as REVAMP was recruiting its last participants.^23^ The general rise in antibody levels to all the antigens evaluated suggests a rise in serology due to community transmission of the virus, rather than through active vaccination campaigns, which only started in March of 2021 and experienced low uptake until the end of 2021.^5^ Notably, the relatively high individual spread of IgG levels observed against SARS-CoV-2 antigens in samples collected before the emergence of SARS-CoV-2—a degree of background reactivity not observed in other cohorts—^17^ and raises questions about potential immunological cross-reactivity between SARS-CoV-2 antigens and other antigens in this population. The clinical significance of this observation remains to be determined.

IgG levels measured against common human coronaviruses as well as Influenza A revealed distinct patterns to those observed against all SARS-CoV-2 antigens, with little variation across the trial period. The stability of Tt antibody levels across all periods served as an important internal control, demonstrating the specificity of the observed changes in antibodies to SARS-CoV-2 and other control viruses. Tt vaccinations were provided by the Government, not as part of the trial, thus demonstrating that despite disruptions to the health services during COVID, ^20^ ANC attendance and services was still relatively strong in this area of Malawi.

Despite the serological evidence of increasing SARS-CoV-2 exposure in the population, during the trial period, pregnancies occurring during COVID-19 experienced longer gestational ages and babies with increased birth weights. As is the case with many respiratory infections,^24^ SARS-CoV-2 infections, and COVID-19 in particular, negatively affect birth outcomes.^25^ However, observations of better outcomes for pregnancy during the pandemic have been made in other cohorts as well.^26^ These results seem to suggest that either 1) exposure levels in our population may not have reached a significant threshold; or 2) the negative impacts of SARS-CoV-2 infection were of less magnitude than the improvements due to reductions in other infections during COVID; or 3) The negative impact of infection is of lesser magnitude than the improvement driven by a shift in the characteristics of the women recruited to the trial— reduced mobility and limited physical demands may have conserved maternal energy, and increased presence of partners at home may have provided added support during pregnancy.

These findings should be interpreted in context. While our study benefits from being embedded within a clinical trial with standardised follow-up and data collection, this also means that our findings may not apply to the broader Malawian population. We did not collect specific information regarding COVID-19 vaccination status of our participants. Our study’s temporal nature makes it challenging to disentangle the effects of SARS-CoV-2 infection from the effects of pandemic-related social and healthcare changes. While our models accounted for demographics such as age and primiparity that shifted during our study, these significant demographic shifts could have introduced unmeasured confounding factors not accounted for in our statistical adjustments. Additionally, it’s important to note that our trial population, receiving regular antenatal care and nutritional supplementation, may have been relatively protected from pandemic-related impacts when compared to the general population^20^.

In conclusion, we demonstrate that in a population of pregnant women in Malawi, where symptoms of COVID-19 were absent and there was no recorded evidence of SARS-CoV-2 infection, population-level IgG to SARS-CoV-2 antigens rose in alignment with well-defined COVID-19 periods. In addition, the increase in inter-antigenic correlation between the antibody levels to SARS-CoV-2 antigens during the COVID-19 period also sustains the interpretation that there was true population transmission during the COVID pandemic. This represents additional evidence to the body of research that proposes pregnancy as a sensitive sentinel period for the population surveillance of infectious diseases. ^11,12,27,28^

## Data Availability

The datasets generated and used during this study are deposited in Figshare under embargo and available after publication using the following link: https://figshare.com/s/1e564308392bbe893bc4

## Author contributions

S-RP and KSP conceptualised and designed the trial and secured funding. EM, GMz, GMh and MNM led data collection in the field. RA and EME planned this study. LMR, NK-W, and KLF conducted experimental data collection. NK-W, RM, EME and RA had full access to and analysed the data. RH, ARDM, SB and RA conducted the statistical analysis. LMR, ARDM, EME and RA interpreted the data. RA prepared the initial draft of the manuscript. All authors critically revised the manuscript for intellectual content, edited and approved the final version, and agreed to be accountable for all aspects of the work.

## Acknowledgements

The Bill & Melinda Gates Foundation funded the REVAMP trial (INV-010612). S-RP is supported by the Australian National Health and Medical Research Council (NHMRC) Fellowships (GNT1158696 and GNT2009047). We thank the local field workers, participants, and their families for their involvement in the study.

## Notes

### Competing Interest Statement

The authors have declared no competing interest.

### Clinical Trial

ACTRN12618001268235

### Author Declarations

The trial received ethics approvals from the College of Medicine, University of Malawi, Malawi (P.02/18/2357), and The Walter and Eliza Hall Institute (WEHI), Australia (18/02), and was prospectively registered (ACTRN12618001268235). An independent data and safety monitoring board oversaw the trial. Secondary analyses of the trial samples were included in the study protocol.

### Summary of Updates

We have revised the title, abstract, and text to highlight the public health significance of the findings. No results or data were changed. We also added a new data availability statement.

