## Supplementary material for "Pregnancy Cohorts as Sentinel Populations: Evidence From Longitudinal SARS-CoV-2 Serology in Malawi"

**Full title:** Pregnancy Cohorts as Sentinel Populations: Evidence From Longitudinal SARS-CoV-2 Serology in Malawi - Supplementary material.

Louise M. Randall, PhD^1*^, Nicholas Kiernan-Walker^1*^, Ernest Moya, PhD^2^, Glory Mzembe, MD^2^, Alistair R. D. McLean, PhD^1,3^, Rebecca Harding, PhD^1^, Ramin Mazhari, PhD^1^, Gomezgani Mhango, MSc^2^, Katherine L. Fielding, MBBS^1^, Ivo Mueller, PhD^1,4^, Martin N. Mwangi, PhD^2,5^, Sabine Braat, MSc^1^, Kamija Phiri, PhD^2^, Sant-Rayn Pasricha, PhD^1,7^, Emily M. Eriksson, PhD^1,4*+^, Ricardo Ataíde, PhD^1*+^

^1^ Infection and Global Health, The Walter and Eliza Hall Institute, VIC, Australia

^2^ Training and Research Unit of Excellence (TRUE), Chichiri, Blantyre, Malawi

^3^ Methods and Implementation Support for Clinical and Health Sciences Research Hub, University of Melbourne, Melbourne, Victoria, Australia

^4^ Department of Medical Biology, University of Melbourne, Melbourne, VIC, Australia

^5^ The Micronutrient Forum, Healthy Mothers Healthy Babies Consortium, Washington, DC, USA.

^6^ Clinical Hematology at the Royal Melbourne Hospital and Peter MacCallum Cancer Centre, Parkville, Australia

^7^ Diagnostic Hematology, The Royal Melbourne Hospital, Parkville, Australia

* These authors contributed equally to the work.

+ Corresponding authors: Ricardo Ataíde,, tel: +61 (03) 9345 2555; Address: The Walter and Eliza Institute for Medical Research, 1G Royal Parade, Parkville, 3052 VIC, Australia, Emily M. Eriksson, tel: +61 (03) 9345 2555; Address: The Walter and Eliza Institute for Medical Research, 1G Royal Parade, Parkville, 3052 VIC, Australia

Supplementary Table 1 - Baseline characteristics according to collection visit

|  | Participants with enrolment samples only | Participants with both timepoints | Participants with delivery samples only |
| --- | --- | --- | --- |
| N (%**)** | 147 (17.3%) | 675 (79.2%) | 30 (3.5%) |
| Maternal age (years) | 20 [18.0, 25.0] | 20 [18.0, 25.0] | 20 [18.0, 24.0] |
| Body mass index (kg/m2)^a^ | 22.9 [21.2, 24.5] | 22.6 [21.0, 24.6] | 22.9 [21.6, 25.1] |
| Primigravid |  |  |  |
| No | 67 (45.6%) | 310 (45.9%) | 14 (46.7%) |
| Yes | 80 (54.4%) | 365 (54.1%) | 16 (53.3%) |
| Religion |  |  |  |
| None | 0 (0.0%) | 1 (0.1%) | 1 (3.3%) |
| Christian | 108 (73.5%) | 483 (71.9%) | 20 (66.7%) |
| Muslim | 37 (25.2%) | 183 (27.2%) | 8 (26.7%) |
| Other | 2 (1.4%) | 5 (0.7%) | 1 (3.3%) |
| Education |  |  |  |
| None | 0 (0.0%) | 2 (0.3%) | 0 (0.0%) |
| Lower primary | 29 (20.4%) | 136 (21.0%) | 7 (23.3%) |
| Upper primary | 58 (40.8%) | 264 (40.7%) | 15 (50.0%) |
| Lower secondary | 11 (7.7%) | 99 (15.3%) | 3 (10.0%) |
| Upper secondary | 40 (28.2%) | 132 (20.4%) | 5 (16.7%) |
| Tertiary | 4 (2.8%) | 15 (2.3%) | 0 (0.0%) |
| Marital status |  |  |  |
| Single | 19 (12.9%) | 110 (16.4%) | 3 (10.0%) |
| Married | 127 (86.4%) | 549 (81.7%) | 26 (86.7%) |
| Widowed | 1 (0.7%) | 2 (0.3%) | 1 (3.3%) |
| Divorced/separated | 0 (0.0%) | 9 (1.3%) | 0 (0.0%) |
| Others | 0 (0.0%) | 2 (0.3%) | 0 (0.0%) |
| Income source |  |  |  |
| None | 5 (3.4%) | 47 (7.0%) | 1 (3.3%) |
| Subsistence farming | 21 (14.3%) | 117 (17.4%) | 9 (30.0%) |
| Large scale farming | 0 (0.0%) | 2 (0.3%) | 0 (0.0%) |
| Employed | 34 (23.1%) | 114 (17.0%) | 4 (13.3%) |
| Casual work for wages | 38 (25.9%) | 213 (31.7%) | 10 (33.3%) |
| Business | 45 (30.6%) | 171 (25.4%) | 6 (20.0%) |
| Other | 4 (2.7%) | 8 (1.2%) | 0 (0.0%) |
| HIV positive |  |  |  |
| No | 121 (83.4%) | 552 (82.4%) | 28 (93.3%) |
| Yes | 24 (16.6%) | 118 (17.6%) | 2 (6.7%) |
| Haemoglobin (g/dL)^b^ | 9.1 [8.0, 10.0] | 8.9 [8.0, 9.6] | 8.7 [8.3, 9.7] |
| Anaemia^c^ |  |  |  |
| No | 8 (5.4%) | 32 (4.8%) | 1 (3.3%) |
| Yes | 139 (94.6%) | 639 (95.2%) | 29 (96.7%) |
| Ferritin (µg/ml) | 28.6 [10.9, 76.1] | 25.7 [9.9, 71.4] | 43.3 [14.9, 74.0] |
| CRP (mg/L)^d^ | 5.4 [3.2, 11.0] | 5.2 [2.7, 10.6] | 5.1 [3.6, 11.9] |
| Inflammation^e^ |  |  |  |
| No | 68 (46.3%) | 328 (49.1%) | 10 (47.6%) |
| Yes | 79 (53.7%) | 340 (50.9%) | 11 (52.4%) |
| Iron deficiency^f^ |  |  |  |
| No | 88 (59.9%) | 371 (55.5%) | 15 (71.4%) |
| Yes | 59 (40.1%) | 297 (44.5%) | 6 (28.6%) |
| Treatment group |  |  |  |
| Oral iron | 74 (50.3%) | 338 (50.1%) | 14 (46.7%) |
| IV iron | 73 (49.7%) | 337 (49.9%) | 16 (53.3%) |

HIV = Human Immunodeficiency Virus; CRP – C-reactive protein

Data are count (%) or median [25% centile, 75% centile]. ^a^ Body Mass Index. ^b^Haemoglobin levels in venous blood, measured by Sysmex. ^c^Anaemia indicates with a venous haemoglobin <11 g/dL. ^d^C-Reactive Protein. ^e^Inflammation indicates a CRP >5 mg/L. ^f^Iron deficient indicates a serum ferritin<15 µg/L, or serum ferritin<30 µg/L if CRP >5mg/L.

**Supplementary Table 2 – Association between time since trial start and level of IgG measured**

|  | **pre-COVID** |  | **Covid wave 1** |  | **Covid wave 2+3** |  |
| --- | --- | --- | --- | --- | --- | --- |
|  | % change over 30 days  [95% Conf. Interval] | P-value | % change over 30 days  [95% Conf. Interval] | P-value | % change over 30 days  [95% Conf. Interval] | P-value |
| **Sars-CoV-2 antigens** |  |  |  |  |  |  |
| S1 | 2.22 [0.16, 4.39] | 0.035 | 0.60 [-1.98, 40.49] | 0.64 | 4.60 [-2.18, 11.96] | 0.19 |
| S2 | -1.49 [-4.11,1.11] | 0.26 | 2.74 [-0.99, 6.61] | 0.15 | 18.53 [8.11, 29.69] | <0.001 |
| RBD | -4.02 [-6.48, -1.58] | 0.0010 | -4.59 [-8.42, -0.56] | 0.026 | 29.69 [18.53, 41.91] | <0.001 |
| Spike | -3.63 [-6.11, -1.09] | 0.0050 | 1.01 [-2.08, 4.39] | 0.50 | 28.40 [17.12, 42.48] | <0.001 |
| NP | -0.70 [-3.05, 1.61] | 0.54 | 1.00 [-4.11, 2.12] | 0.52 | 22.02 [12.19, 32.71] | <0.001 |
| **Seasonal Viruses** |  |  |  |  |  |  |
| Coronavirus 229e | -2.96 [-6.57, 0.60] | 0.10 | 0.36 [-4.37, 5.34] | 0.88 | -5.14 [-13.50, 4.08] | 0.27 |
| Coronavirus HKU1 | -4.11 [-5.82, -2.33] | <0.001 | 6.82 [4.24, 9.53] | <0.001 | -4.88 [-10.77, 1.31] | 0.12 |
| Coronavirus OC43 | 0.53 [-1.39, 2.53] | 0.59 | 4.60 [1.71, 7.68] | 0.0020 | -6.29 [-12.80, 0.73] | 0.078 |
| Coronavirus NL63 | -1.64 [-3.83, 0.61] | 0.15 | 5.13 [2.02, 8.33] | 0.0010 | -7.32 [-13.50, -0.83] | 0.028 |
| Influenza A (H1N1) | -1.69 [-4.18, 0.85] | 0.19 | 5.44 [1.64, 9.50] | 0.0050 | -6.11 [-13.76, 2.22] | 0.15 |

Notes: Outcome variables were log_e_-transformed before running the models. Associations were sought with spline regression fitting models with three segments (up to April 2nd 2020, between April 2nd 2020 and January 1st 2021 and after January 1st 2021). Multivariable models included maternal age, body mass index, primiparity, sex of the newborn, HIV status, income source and treatment group. Multivariable models were run on all women with delivery samples available with non-missing data for all variables (N=544).

Supplementary Table 3 – Associations between pregnancy outcomes and seropositivity to SARS-CoV-2 antigens

| Exposure: sars-cov-2 seropositivity inside COVID period | Mean difference or Odds Ratio  [95% Conf. Interval] | P-value |
| --- | --- | --- |
| Outcome: gestational age (weeks) | 0.07 [-0.5, 0.6] | 0.79 |
| Outcome: birth weight (grams) | 79.7 [-93.3, 252.7] | 0.37 |
| Outcome: low birth weight (<2500 grams) | 0.88 [0.34, 2.30] | 0.80 |

Note: Multivariable models included maternal age, body mass index, primiparity, sex of the newborn, HIV status, income source and treatment group. Multivariable models were run on all women whose pregnancies occurred within the COVID period and with non-missing data for all variables (Gestational age, N = 248; Birth weight, N = 244; Low birth weight, N = 244).

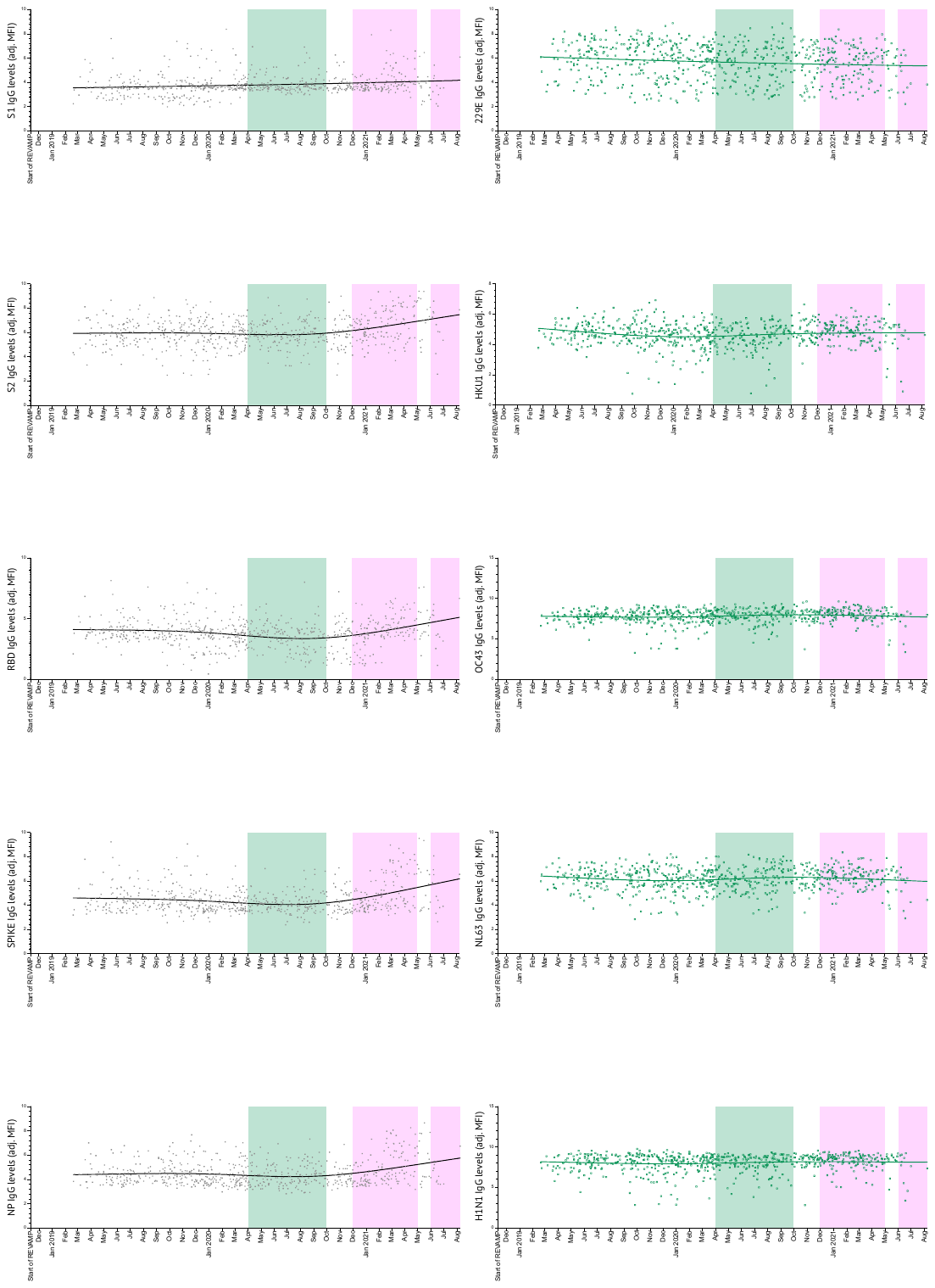

**Supplementary figure 1 - Serological patterns of anti-SARS-CoV-2** **antibodies during pregnancy.** LOWESS smoothing curves illustrating the relationship between time and anti-SARS-CoV-2 antibodies is shown for all SARS-CoV-2 (left panels) and all control antigens (right panels). This method fits a non-parametric regression and makes no assumptions about the functional form of the relationship. The graph includes the original data points as scatter markers to allow assessment of the density and distribution of observations. Shaded boxes represent reported COVID waves (wave 1 – green; waves 2 and wave 3 – pink).
